# A novel splice site variant in *DEGS1* leads to aberrant splicing and loss of DEGS1 enzyme activity, a VUS resolved

**DOI:** 10.1101/2025.04.04.25325118

**Authors:** Holly C. Beale, Victor Tse, Joanna Y. Lee, Jon Akutagawa, Yusuph Mavura, Brandon Saint-John, Allison Cheney, Dennis R. Mulligan, Guillermo Chacaltana, Martin Gutierrez, Jessica Tenney, Joseph T. Shieh, Pierre-Marie Martin, Tiffany Yip, Ugur Hodoglugil, Alex J. Fay, Angela N. Brooks, Jessica Van Ziffle, Michael D. Stone, Neil Risch, Jeremy R. Sanford, Patrick Devine, Julie D. Saba, Olena M. Vaske, Anne Slavotinek

## Abstract

**Purpose:** Pathogenic *DEGS1* variants have been reported in individuals with autosomal recessive hypomyelinating leukodystrophy 18 (HLD18; MIM# 618404). We sought to resolve a 5′ +4/+5 splice site variant of uncertain significance found in three individuals with HLD features.

**Methods:** We used next-generation DNA and transcriptome sequencing, cell-based splicing assays, and tandem mass spectrometry to detect and characterize the splice site variant. We then performed RNA structure probing and conventional antisense oligonucleotide screening to investigate molecular mechanisms for potential therapeutic intervention.

**Results:** A homozygous, *DEGS1* 5′ splice site variant, c.825+4_825+5delAGinsTT (NM_003676.4) was identified in all three participants. Although the gene has been associated with autosomal recessive hypomyelinating leukodystrophy, the variant has not been previously reported in any available databases or literature. We show that the splice site variant: 1) was sufficient to induce exon two skipping in most detected transcripts; 2) resulted in structural changes to the 5′ and 3′ splice site regions using RNA structure probing; and 3) corresponds to plasma sphingolipid profiles consistent with loss of sphingolipid delta(4)-desaturase activity.

**Discussion:** Our RNA and lipidomic evidence proved that the *DEGS1* variant c.825+4_825+5delAGinsTT is pathogenic and suggested a mechanistic model to explain how exon two skipping is induced.

## Introduction

Hypomyelinating leukodystrophies (HLD) are a heterogeneous group of heritable, progressive disorders in which the formation of myelin sheaths is disrupted, resulting in neurodegenerative white matter disease that can affect the function of neurons and impair cognitive and motor skills.^1–3^ Clinical manifestations of HLD are highly variable, even for individuals within the same family, but often include motor regression and neurological findings such as dystonia, spasticity, abnormal tone, developmental delay, seizure, nystagmus, cognition and learning impairment.^3–5^ Currently, twenty-seven genes are associated with HLD in the Online Mendelian Inheritance in Man database (MIM# 312080).

Loss of function variants in *Delta 4-Desaturase, Sphingolipid 1* (*DEGS1*) are associated with autosomal recessive hypomyelinating leukodystrophy 18 (HLD18; MIM# 618404). Approximately thirty individuals with HLD18 have been reported in the literature to date.^4,6–11^ *DEGS1* is a three exon gene that functions in the *de novo* ceramide biosynthesis pathway. The product of the *DEGS1* gene, DES1, is an enzyme that catalyzes the conversion of dihydroceramide (DHCer) to ceramide (Cer) through the introduction of a double bond into the backbone of the sphingolipid.^12^ The conversion of saturated to unsaturated sphingolipids is an essential process that maintains the myelin that surrounds and protects the axons of neurons.^13^

Aberrant splicing of precursor messenger RNA (pre-mRNA) is a common defect in many inherited diseases.^14,15^ These gene expression errors typically arise from variants at the consensus splice site sequences that define exon-intron boundaries. Accurate expression of the *DEGS1* gene requires removal of two introns from the pre-mRNA. To our knowledge, no publications report splice site variants in HLD18 patients. However, the ClinVar database contains three *DEGS1* splice site variants: Variation ID #2502429 is likely pathogenic and found in an affected individual; #1514221 is likely pathogenic in an unaffected individual; and #1481460 is of uncertain significance found in an unaffected individual (Supplemental Table 1). These results suggest a potential role for aberrant splicing in HLD.

In this study, we report the discovery and characterization of a *DEGS1* splice site variant found in three participants with clinical features consistent with HLD18 from two unrelated families. We then performed RNA structure probing and conventional antisense oligonucleotide (ASOs) screening to investigate molecular mechanisms for therapeutic intervention. Our results provide a mechanistic understanding of this *DEGS1* pathogenic variant and raise the intriguing possibility that ASOs may provide a novel therapeutic option for some HLD patients.

## Materials and Methods

Written, informed consent was obtained from the parents of participants one, two and three using a consent form from the [REDACTED], part of the [REDACTED], that was approved by the Institutional Review Board (IRB) at [REDACTED].

### Exome sequencing and variant identification

Exome sequencing was performed as previously described on DNA isolated from blood or saliva submitted by participants and family members, respectively.^16^ Briefly, targeted regions were selected with the xGen Whole Exome Panel kit v1 (Integrated DNA Technologies) and sequenced using the Illumina HiSeq 2500 sequencing system with v3 chemistry generating 100bp paired-end reads in rapid run mode. Mean depths of coverage for participants one and two were 94X reads and 102X reads, and 99.5% and 99.6% of exons, respectively, had at least 10X coverage. The resulting DNA sequences were mapped to the human genome GRCh37/hg19.

Sequence variant detection, filtering, ranking and annotation was performed using Opal Clinical (Fabric Genomics) and Moon Diploid. The variants found in the participants were compared to variants in other family members to annotate the variants based on their inheritance patterns: *de novo*, homozygous, compound heterozygous and inherited heterozygous. The inherited heterozygous variants were filtered based on the following key words and Human Phenotype Ontology (HPO) terms. Key word(s) and/or HPO terms used to filter and rank variants in the first participant included: severe global developmental delay (HP:0011344), contractures (HP:0001371), seizures (HP:0001250), generalized hypotonia (HP:0001290), cortical visual impairment (HP:0100704), optic atrophy (HP:0000648). Key word(s) and/or HPO terms used to filter and rank variants in the second participant included: seizures (HP:0001250), global developmental delay (HP:0001263), hypertonia (HP:0001276), microcephaly (HP:0000252), spastic quadriparesis (HP:0001285), cerebral palsy (HP:0100021), facial dysmorphism (HP:0001999), hirsutism (HP:0001007), cerebellar atrophy (HP:0001272), white matter abnormalities (HP:0002500), hypomyelination (HP:0003429). The variant data were also analyzed using a manual pipeline whereby bioinformatics experts from [REDACTED] manually curated the inherited heterozygous variants with the gene lists and keywords in Supplemental Table 2. In the third participant, who was the sibling to the second patient, DNA from whole blood was analyzed using a Leukodystrophy and Genetic Leukoencephalopathy Panel (Invitae, Inc.).

We identified a splice site variant in *DEGS1* which replaced the nucleotides AG with TT at position 4 of the exon/intron boundary (NM_003676.4) c.825+4_825+5delAGinsTT in the exome sequencing. This variant was confirmed to be homozygous in participants one and two by Sanger sequencing. Briefly, amplification of genomic DNA with forward primer ACCGATTTTGAGGGCTGGTT and reverse primer ATGAACTGCTTGGACTGACA, was followed by sequencing with a BigDye Terminator 3.1 Cycle Sequencing Kit and Genetic Analyzer 3500 (ThermoFisher Scientific).

### Genetic relatedness estimation using PC-Relate

We estimated the relatedness of participants and parents from two families using the program PC-Relate^17^ as previously described using six principal components.^18^

### Transcriptome sequencing and isoform analysis

RNA was extracted from whole blood (Maxwell® RSC simplyRNA Blood Kit, Promega catalog # AS1380) from participant one and an unrelated control without any pathogenic variants or a variant of uncertain significance (VUS) in *DEGS1*. Libraries were prepared with the KAPA Stranded mRNA-Seq Kit with KAPA mRNA Capture Beads (Roche catalog #07962193001). 150-base paired end sequence was generated from libraries on an Illumina Novaseq 6000 sequencer using an S4 flow cell. Participant one data had 82.6 million total pairs of reads, of which 45.1 million were Mapped Exonic Non-Duplicate (MEND) reads.^19^ The control data had 87.7 and 22.9 million total and MEND reads, respectively. For comparison to reference genome and transcripts, reads were aligned by STAR v2.4.2a using indices generated from the human reference genome GRCh38 and the human gene models GENCODE 23. To assemble novel transcripts, hisat2:2.2.1 was used to align reads to GRCh38 (with the index grch38_snp_tran.tar.gz downloaded from ftp://ftp.ccb.jhu.edu/pub/infphilo/hisat2/data/). Novel and reference transcripts were identified using Stringtie 2.1.6, with and without Gencode v38 as a guide. Additionally, Stringtie was run on aligned reads after duplicates were removed with samblaster 0.1.26. All outputs were merged to generate a gene transfer format (GTF) file with consistent identifiers containing all reference and de novo transcripts, and transcript quantification was repeated with Stringtie using the merged GTF as a guide. The fraction of *DEGS1* expression accounted for by each isoform was calculated based on the TPM of each isoform. We report isoform abundance using duplicate-free data; the abundances based on data inclusive of duplicate reads are similar.

### *DEGS1* Junction Usage Analysis

Percent-spliced (PS) values for splice junctions were computed with mesa v1.0.0 (https://github.com/BrooksLabUCSC/mesa) for participant one, the RNA-Seq control, and 670 whole blood samples from the Genotype-Tissue Expression (GTEx) v8 dataset^20^. Reads were aligned to the hg38 reference genome using STAR 2.4.2a. A PS value for a splice junction (inclusion) is calculated by the total read count for that junction, divided by the total of the inclusion and all exclusion junctions. The exclusion are reads containing any other splice junction that overlaps the inclusion splice interval that is being quantified. These intervals are considered mutually exclusive, and represent some form of alternative splicing. A distribution of PS values from the GTEx whole blood samples was used to calculate the quartile ranges.

Outlier splicing events were defined as events that were larger than the third quartile (Q3) + 1.5 * the interquartile range (IQR) or smaller than the first quartile (Q1) - 1.5 * IQR. PS values from participant one and the RNA-Seq control were then compared to these cutoffs.

### *DEGS1* Splicing Reporters

Reference and variant *DEGS1* exon two splicing reporters were generated and validated as previously described.^21^

### Cell-based In Vivo Splicing Reporter Assays

HEK293T cells (ATCC) were cultured in 6-well tissue culture plates (CytoOne, USA Scientific) using Dulbecco’s Modified Eagle Medium (Gibco™, supplemented with 10% FBS) at 37°C with a CO_2_ level of 5%. Prior to the time of performing the assays, cells were grown to a confluency of ∼60-80%. 2.5 µg of each splicing reporter was transiently transfected into HEK293T cells using lipofection technology (Lipofectamine 2000). At 24 hours post transfection, cells were harvested and prepared for total RNA purification using the Direct-zol RNA Miniprep kits from Zymo Research.

### Antisense Oligonucleotides (ASOs)

*DEGS1* exon two ASOs were designed by taking the reverse complement of the coding sequence, specifying sequences of *k*-mer length, which were then annotated with desired modifications to the ribose sugar. To infer nuclease resistance and *in vivo* stability to ASOs, the 2’OH contained a methoxyethyl modification (2’MOE) and the phosphate backbone was modified to a phosphorothioate backbone. *DEGS1* ASOs were designed to be 18 nucleotides in length and were synthesized by Integrated DNA Technologies (IDT).

### *DEGS1* Reference and Variant ASO Walks

HEK293T cells (ATCC) were cultured in 96-well tissue culture plates (Perkin Elmer) using Dulbecco’s Modified Eagle Medium (Gibco, supplemented with 10% FBS) at 37°C with a CO_2_ level of 5%. Prior to performing the assays, cells were grown to a cell confluency of ∼60-80%. 250 ng of reference or variant *DEGS1* splicing reporters were transiently co-transfected with 10 pmol of each ASO into HEK293T cells using lipofection technology (Lipofectamine 2000). After 24-hours post transfection, cells were then harvested and prepared for total RNA purification using the *Quick*-DNA/RNA Viral MagBead kit from Zymo Research, in which this workflow has been automated on the Agilent Bravo.

### 2-step RT-qPCR and Qualitative Analysis of Splicing Reporter Assays

First-strand cDNA synthesis of total RNA using Multiscribe Reverse Transcriptase (Applied Biosystems) and 5’FAM end-labeled PCR amplification of mRNA reporter isoforms was performed following protocols as described.^21^ The resulting amplicons were then analyzed using gel electrophoresis to empirically evaluate mRNA isoforms detected.

### Quantitative Analysis of Splicing Reporter Assays using Fragment Analysis

The abundance of each 5’FAM end-labeled amplicon was quantified and analyzed following protocols as described.^21^

### Calculating Splicing Efficiency using Percent-Spliced-In (PSI) Index Formula

Quantification of reference or variant *DEGS1* exon two splicing efficiency was determined following protocols as described.^21^

### *DEGS1* Reference and Variant Exon Two RNA synthesis

*DEGS1* exon two RNA(s) were synthesized by T7 RNA polymerase using a *DEGS1* reference/variant construct containing intron spanning regions as a template. RNA was purified using standard agarose gel extraction followed by overnight ethanol precipitation.

### In-vitro Mutational Profiling coupled with High Throughput Sequencing (MaP-seq)

MaP-seq experiments were performed and data were analyzed with RNAFramework and RNAstructure as previously described.^21^

### Human subjects for healthy control blood collection

Pediatric plasma samples from nine healthy children and young adults were obtained from the [REDACTED] in a deidentified state after selection by the clinical laboratory manager. Blood was collected from participants in accordance with an approved IRB protocol at [REDACTED]. Inclusion criteria were male and female pediatric subjects 0 - 20 years of age who were undergoing preoperative lab testing prior to elective surgery. Exclusion criteria were any subjects with a metabolic, malignant, infectious, autoimmune or hemolytic diagnosis. Samples were maintained at 4°C and processed within 24 hours of the time of collection.

### Plasma isolation

Samples of EDTA chelated whole venous blood were processed by centrifugation at 4°C at 350 x g for five minutes. Supernatant was transferred to a new tube and centrifuged at 8,050 x g at 4°C for five minutes. The clear plasma was transferred to new microcentrifuge tubes, snap frozen with dry ice, and stored at −80°C until lipid extraction. Hemolyzed blood samples were excluded from the analysis.

### Liquid Chromatography Mass Spectrometry (LC-MS/MS) Detection of Sphingolipids

Plasma sphingolipids were extracted as previously described.^22^ Detection of sphingolipid and data processing methods were used as previously described.^23^ Data processing was performed using the Agilent quantitative analysis application. All samples were analyzed using triplicates. All data are expressed as the mean ± standard deviation (SD). Differences were examined for significance using the two-tailed Student’s test (t test), with p < 0.05 as the cutoff for statistical significance.

## Results

### Clinical presentation

We present three participants from two families. All participants had severe motor delays, with failure to achieve independent sitting by [REDACTED] of age (participant three), [REDACTED] years (participant one), and [REDACTED] years (participant two). All three participants were noted to have nystagmus in the neonatal period. All had feeding difficulties with resultant failure to thrive, microcephaly, abnormal tone, and weakness. None were able to babble or acquire single words. Participants one and two manifested seizures and developed limb contractures involving the wrists and knees and, in one, the elbows. Participant one (Figure 1A) developed respiratory failure and was deceased at [REDACTED] years [REDACTED] months, whereas participant two suffered respiratory decline at [REDACTED] years of age and required a tracheostomy at [REDACTED] years. In participant one, magnetic resonance imaging (MRI) of the brain showed abnormal myelination development (Figure 1B). For a full description of clinical findings of these participants see Supplemental Table 3.

**Figure 1.**
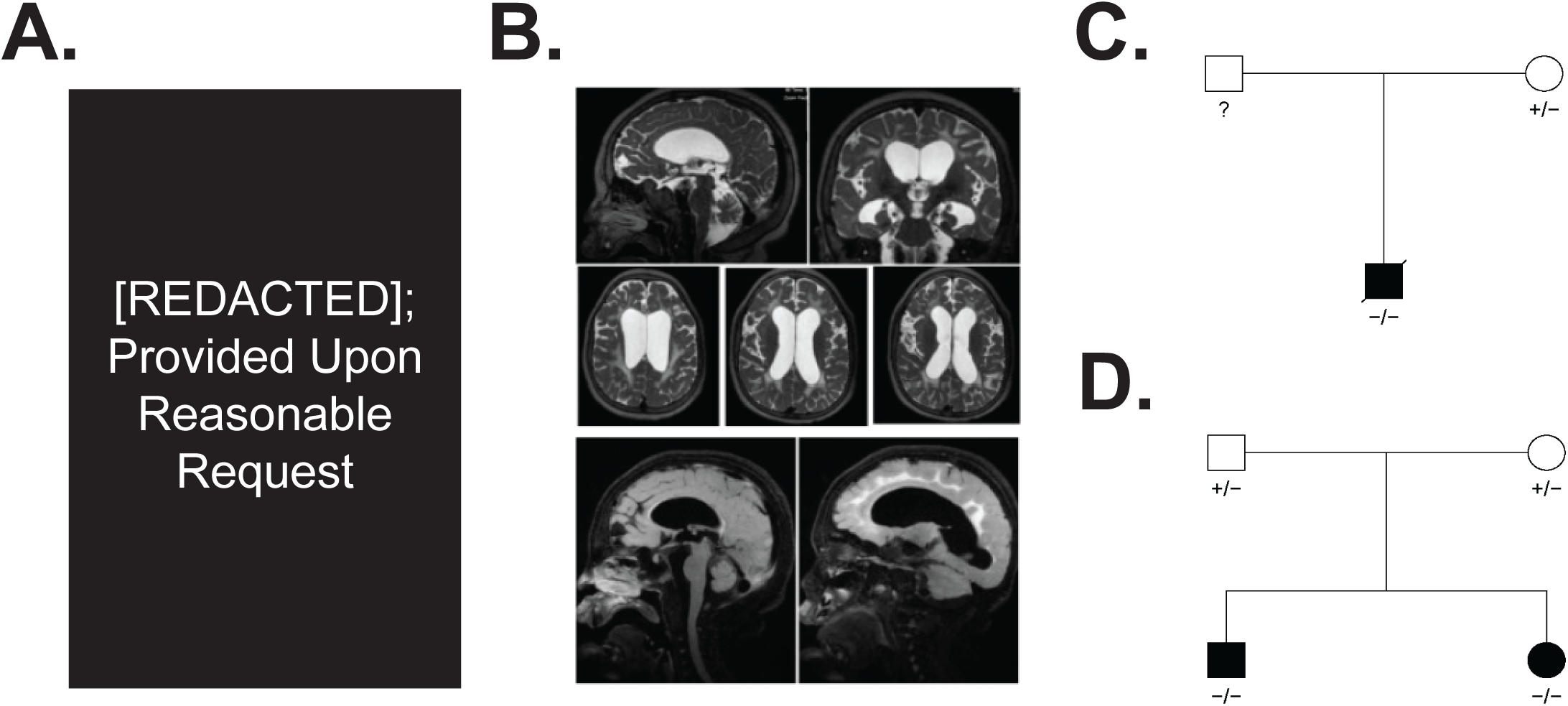
Clinical presentation of unrelated patients with a previously unreported homozygous, 5’ splice site variant in *DEGS1*. **(A)** Facial photograph of the first participant, showing prominent eyes with partial ptosis, a depressed nasal bridge, anteverted nares, long philtrum, broad and wide mouth and micrognathia. He has thick eyebrows and long eyelashes. **(B)** Magnetic resonance imaging (MRI) of the brain for participant one showed profound paucity white matter, thinning of the corpus callosum, volume loss of the midbrain and vermis, and periventricular heterotopia of the left frontal horn. **(C)** Pedigree of family one and participant one. **(D)** Pedigree of family two with participant two (male) and participant three (female).

### Detection of a splice site variant of uncertain significance in *DEGS1*

All participants shared a homozygous *DEGS1* (NM_003676.4) 5’ splice site variant (Figure 1C, D), NM_003676.4:c.825+4_825+5delAGinsTT, which replaced the nucleotides AG with TT at position 4 and 5 of the exon/intron boundary (Figure 2A; NC_000001.10:g.224378025_224378026delinsTT (GRCh37)). The variant was heterozygous in all available parents (Supplemental Figure 1A, B).

**Figure 2.**
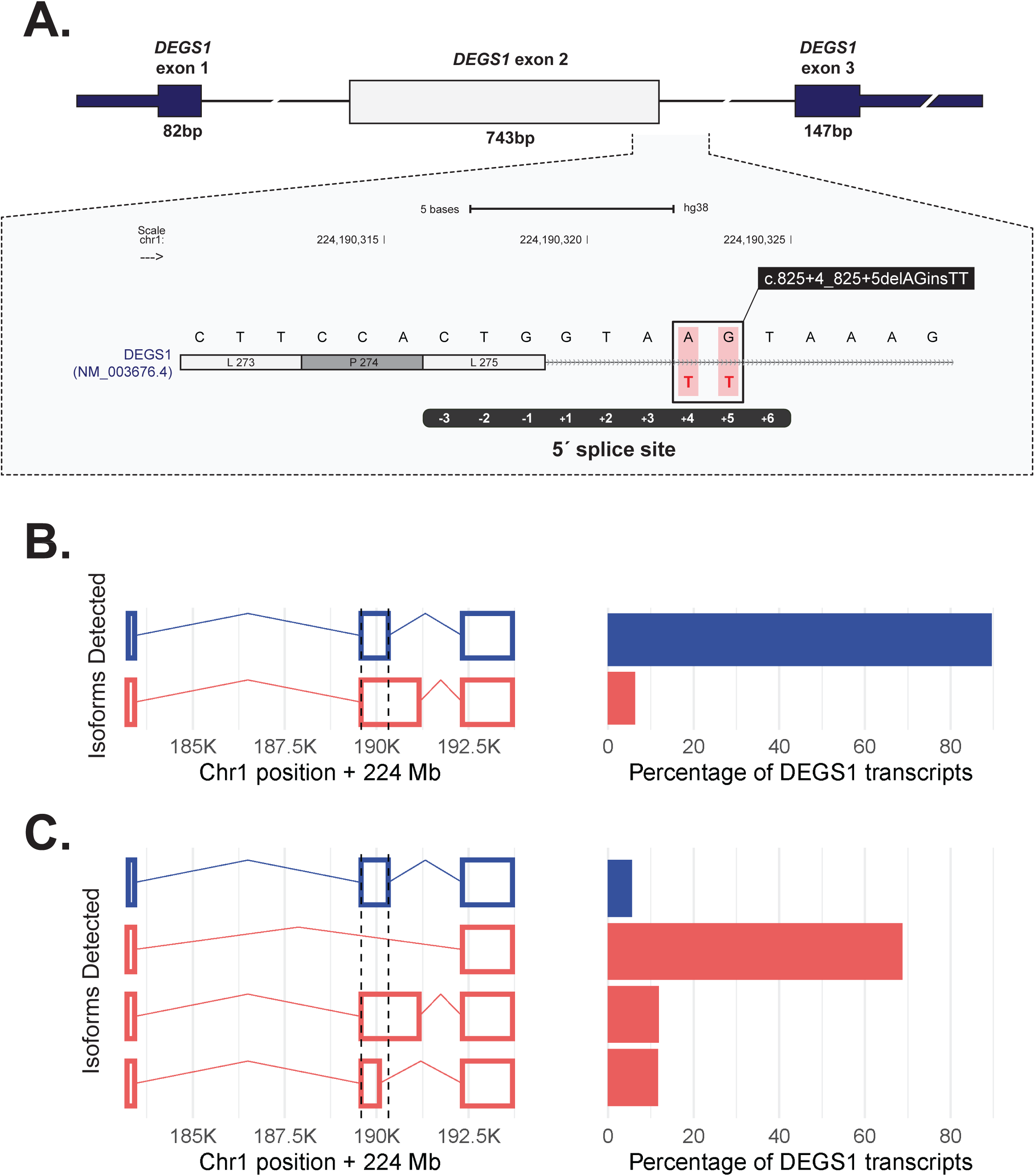
*DEGS1* variant and isoforms detected. **(A)** The c.825+4_825+5delAGinsTT variant maps to the 5′ splice site of exon two. The position of the variant is boxed in the enlarged inset. **(B)** *DEGS1* isoforms (left) present at more than 5% in the control and the corresponding abundance (right). The isoform plots depict canonical (blue) and non-canonical isoforms (red). The canonical transcript comprised most of the transcript molecules. **(C)** *DEGS1* isoforms and abundance in participant one. Transcript assembly identified two non-canonical transcripts not detected in the control. The novel transcript that skips exon two is the predominant transcript.

This variant was annotated as a variant of uncertain significance (VUS) because there was no sufficient evidence from prior reports or functional studies to infer pathogenicity. This classification was based on the American College of Medical Genetics and Genomics (ACMG) guidelines.^24^ Specifically, the variant was classified as a VUS as it was: (1) absent from population databases (e.g., gnomAD, dbSNP, TOPMed, DiscovEHR)^25–28^ and (2) has not previously been published in the literature or associated with disease in databases such as ClinVar. Both families self-reported their ancestry as [REDACTED], suggesting a potential founder mutation in that ancestry.^29^ We also determined that the two families are only distantly related, at least to the fourth degree (Supplemental Figure 2). However, we also noted that the splice site variant was carried on a shared haplotype extending thousands of nucleotides between the three patients and two families, confirming a common founder origin.

### Exon two of *DEGS1* is skipped in transcripts from participant one

Two splice effect prediction algorithms indicated the variant would have a deleterious effect on splice sites. SpliceAI^30^ reported a 94% probability of donor loss (https://spliceailookup.broadinstitute.org, accessed 3/4/2025), and MaxEntScan^31^ reported reduced strength score for the 5’ splice site from 10.65 for the wild type sequence to 3.5 for the variant sequence (Supplemental Table 4).

To determine whether the splice site variant had a functional impact on *DEGS1* splicing, we compared transcripts in whole blood from an individual without pathogenic or uncertain variants in *DEGS1* (the control), participant one, and GTEx, which is a large cohort of normal samples. We first quantified *DEGS1* isoforms. In whole blood in GTEx, the MANE select transcript ENST00000323699 (NM_003676) comprised 97.6% of expressed reference transcripts (GTEx transcript browser https://www.gtexportal.org/home/transcript, page accessed 02/11/2025). When we assembled *DEGS1* transcripts from RNA sequencing data, 90% of *DEGS1* transcripts detected in the control sample were the reference transcript NM_003676 (Figure 2B). In contrast, 69% of *DEGS1* transcripts detected in participant one corresponded to a novel transcript that differed from NM_003676 only by skipping the second of three exons (Figure 2C; Supplemental Tables 5 and 6). This novel isoform is predicted to correspond to the amino acid change NP_003667.1:Ala28Glyfs*7.

We next quantified splice junction usage. As one would expect from the prevalence of the three-exon NM_003676 in normal blood, evidence for usage of exon one-two junctions and exon two-three junctions was abundant in the control and GTEx whole blood data. In participant one data, usage of those exons fell below the down-outlier threshold (Supplemental Figure 3). The exon one-three junction, which is not present in reference gene models, was abundant in participant one and was not found in the control. In GTEx whole blood, only 44% of samples used the *DEGS1* exon one-three junction at all, and at most, that junction usage comprised 10% of *DEGS1* junction usage. In comparison, it comprised 56% of junction usage in participant 1 (Supplemental Table 7).

### The splice site variant is sufficient to induce complete skipping of *DEGS1* exon two

To experimentally validate computational predictions and our RNA-seq data, we performed cell-based splicing reporter assays to examine *DEGS1* exon two inclusion in the reference and variant contexts (Figure 3A). Relative to the reference (lanes 1-3), the splice site variant induced complete and significant skipping of *DEGS1* exon two from the splicing reporter (lanes 6-8) (Figure 3B, C). This finding recapitulated and supported the predominant exon two skipping isoform detected from RNA-seq of participant one (Figure 2C). Together, these findings show that this novel splice site variant was sufficient by itself to cause significant skipping of *DEGS1* exon two.

**Figure 3.**
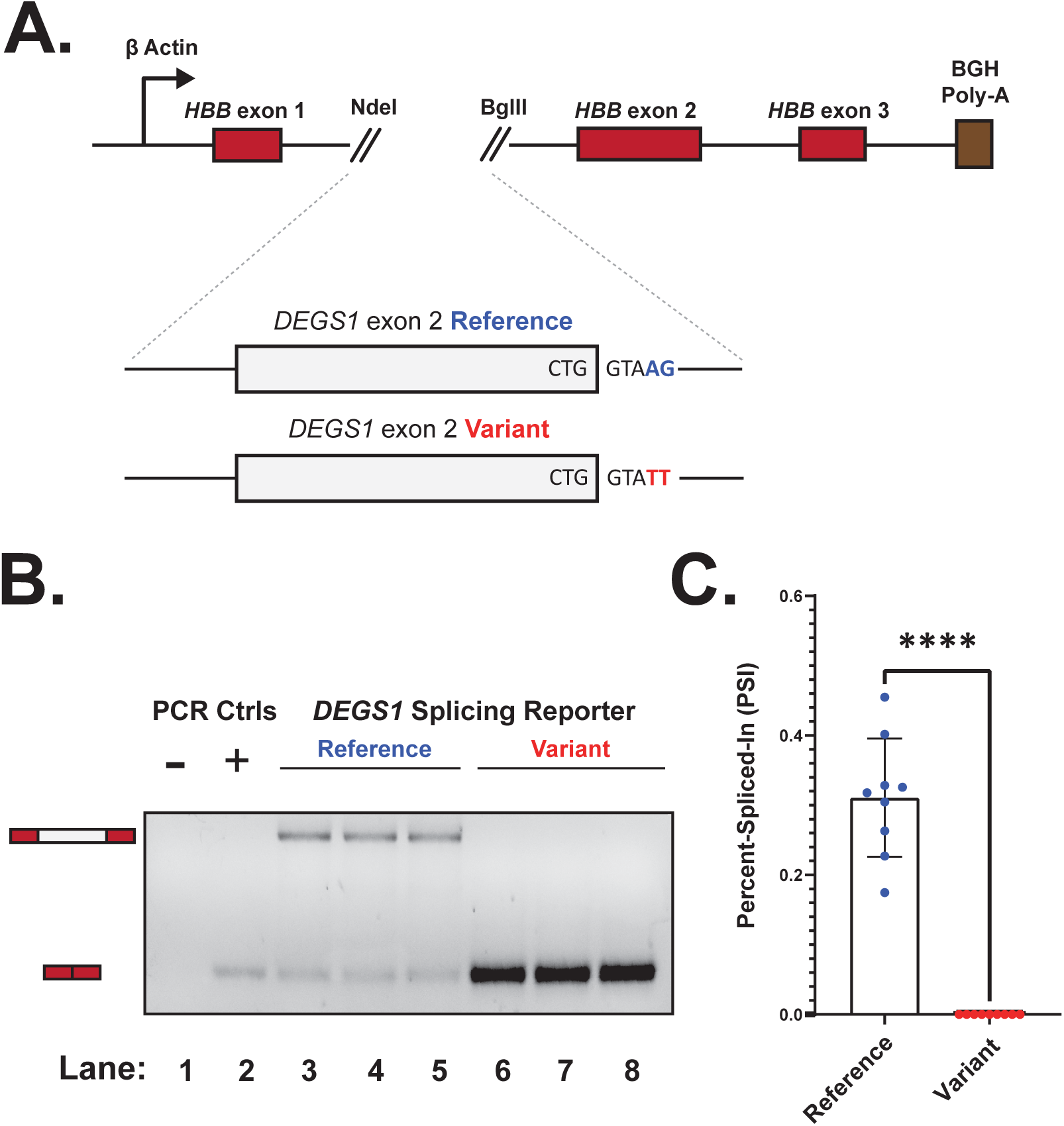
Splice site variant is sufficient to induce skipping of *DEGS1* exon two. **(A)** Schematic depicting the heterologous splicing reporter system used to assay the functional impact the novel splice site variant has on *DEGS1* exon two splicing, relative to the reference. **(B)** A representative agarose gel showing the variant’s effect on *DEGS1* exon two splicing. As shown in the annotation above the representative agarose gel, controls include a no template reaction (lane 1) and a positive control for exon skipping (lane 2). Lanes corresponding to the splicing reporters that assayed the splice site variant or reference sequence context of *DEGS1* exon two are indicated respectively. Expected mRNA isoforms including or excluding the *DEGS1* exon two are also annotated to the left of the agarose gel. **(C)** Percent-spliced-in (PSI) plot quantifying the results shown in **(B)**, measuring the splice site variant’s impact on *DEGS1* exon two inclusion. PSI refers to the fraction of mRNA reporter isoforms that include the exon of interest, relative to the total population of mRNA reporter isoforms. Statistical significance between comparisons shown is denoted by asterisks (i.e., ****) that represent a *P* ≤ 0.0001. Statistical significance was determined using analysis of variance (ANOVA), and Dunett’s post-hoc test. Each condition tested and presented contains nine independent/biological replicates.

### Splice site variant drives substantial refolding of RNA secondary structures

Since RNA structure formation has been implicated to have clinical relevance,^32^ we investigated the potential for *DEGS1* exon two, an unusually large exon, to form local and long-range RNA secondary structures. To test the hypothesis that this splice site variant can affect the RNA folding landscape of *DEGS1* exon two, we used SHAPE-MaP-seq (<u>s</u>elective 2’-<u>h</u>ydroxyl <u>a</u>cylation analyzed by <u>p</u>rimer <u>e</u>xtension and <u>m</u>ut<u>a</u>tional <u>p</u>rofiling coupled to high-throughput sequencing) to chemically probe the accessibility of the reference and variant pre-mRNA context corresponding to *DEGS1* exon two. SHAPE-MaP-seq revealed striking differences between the RNA structure profiles of the reference and variant (Figure 4A, compare blue to red; Supplemental Figure 4; Supplemental Figure 5). In addition to further weakening of the 5′ splice site by the variant through its sequestration in a long-range structure, we also saw that the 3′ splice site in the variant context is now structured, instead of being in an apical loop as seen in the reference (Supplemental Figure 6). A conventional ASO walk, tiling chemically-modified ASOs across a gene target,^33^ did not identify any splice-modulating ASOs for the pathogenic variant, but the same walk on the reference sequence revealed striking splicing inhibition by multiple ASOs (Supplemental Figure 7). The sequence of successful inhibitory ASOs overlapped with putative binding sites for splicing enhancing factors, as indicated by our RBPmap analysis (Supplemental Figure 8).^34^ Together, our RNA folding models suggest this splice site variant rearranges the RNA structure profile of *DEGS1* exon two (Figure 4B, C), weakening its accessibility between: 1) U1 snRNA and the 5′ splice site; 2) U2AF heterodimer and the 3′ splice site; 3) and possibly splicing factors to regulatory sequences as indicated by our ASO walk.

**Figure 4.**
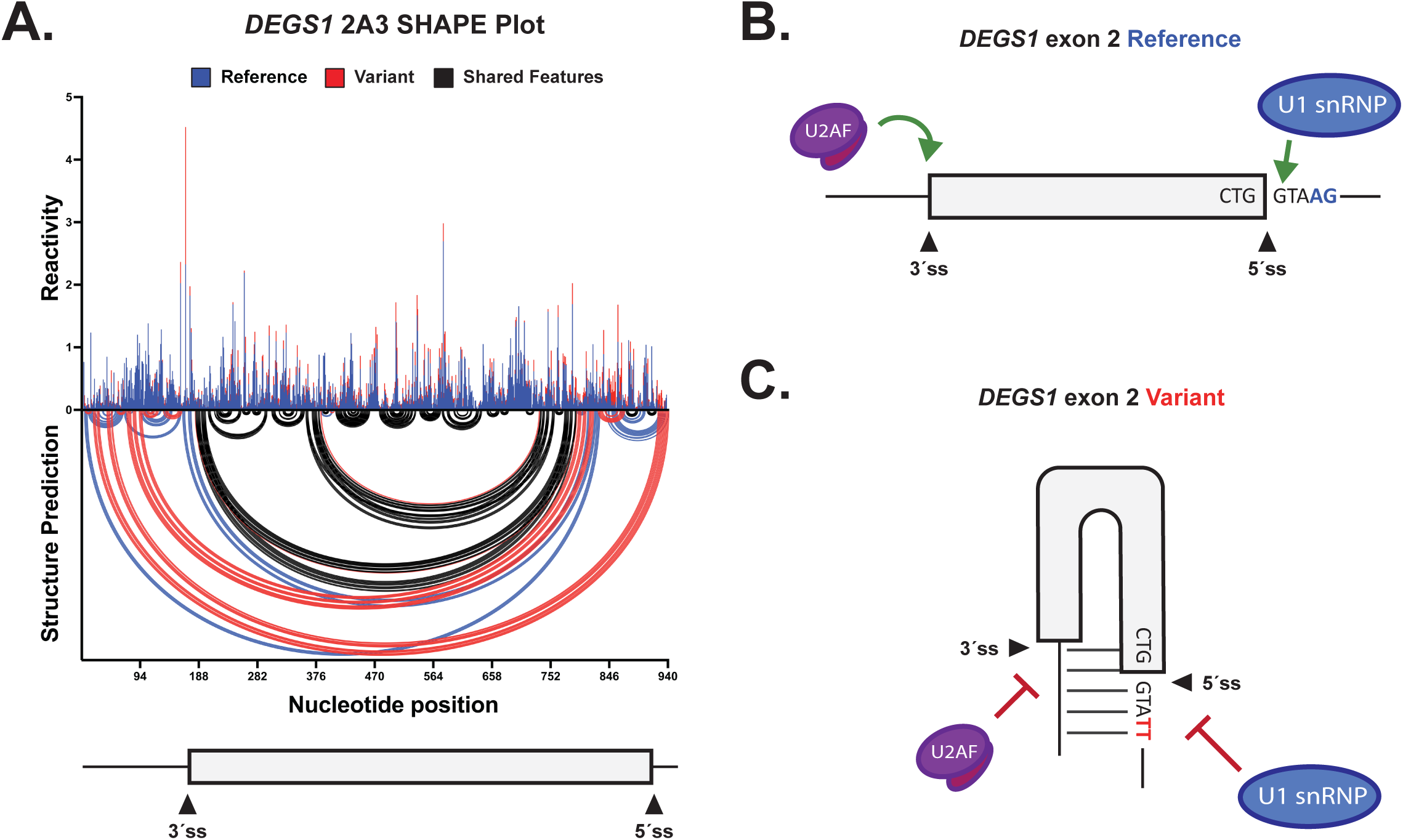
Splice site variant drives intramolecular refolding of RNA secondary structures of *DEGS1* exon two. **(A)** A normalized SHAPE reactivity versus structure prediction plot comparing the RNA folding profiles for the reference and variant sequence context of *DEGS1* exon two (depicted in blue and red, respectively). The top part of the plot shows the normalized SHAPE reactivity for each nucleotide. The bottom part of the plot shows SHAPE-constrained structure predictions represented by intramolecular base pairing interactions; secondary structure formation is denoted by arcs joining different regions of the RNA sequence context. A schematic model of *DEGS1* exon two and its flanking introns is also shown at the bottom of the plot to illustrate relative positions of RNA structure data. All SHAPE data analysis was performed in RNA Framework. **(B, C)** Simplified models of *DEGS1* exon two in the reference and splice site variant context. Critical spliceosomal components and position of splice sites are indicated. Base pairing is indicated by horizontal solid lines between regions, as seen in (C).

### Reduced desaturase activity observed in the tested probands

Because the DES1 protein encoded by *DEGS1* catalyzes the insertion of a double bond into the backbone of sphingolipids during ceramide synthesis (Fig 5A), we used tandem mass spectrometry to quantify the saturated and unsaturated sphingolipids in plasma samples from participant one and participant two and compared the results to nine pediatric controls. An overall profile of high DHCer levels, low Cer levels, and high DHCer/Cer ratios was evident in both participants compared to controls (Supplemental Table 8; Supplemental Figure 9). For most lipid species, Cer levels were lower in probands than the average control level. These changes were profound and present across a broad range of DHCer and Cer species, which vary based on the chain length and saturation of the fatty acid component of the sphingolipid. For example, the ratios of C14:0, C16:0, C20:0, C22, C24:0, C24:1 and C26:0 DHCer/Cer were elevated from 42-fold to 44,300-fold in the probands compared to controls (Figure 5B)., and Cer was significantly lower in participants than controls for C14:0, C16:0, C18:0, C20:0, C22:0, C24:0, C24:1, and C26:0 (Supplemental Figure 9). These measurements confirmed a functional deficiency of ceramide desaturase activity. Together, these lipidomic analyses demonstrate that this splice site variant of *DEGS1* leads to a loss-of-function of the protein product, DES1.

**Figure 5.**
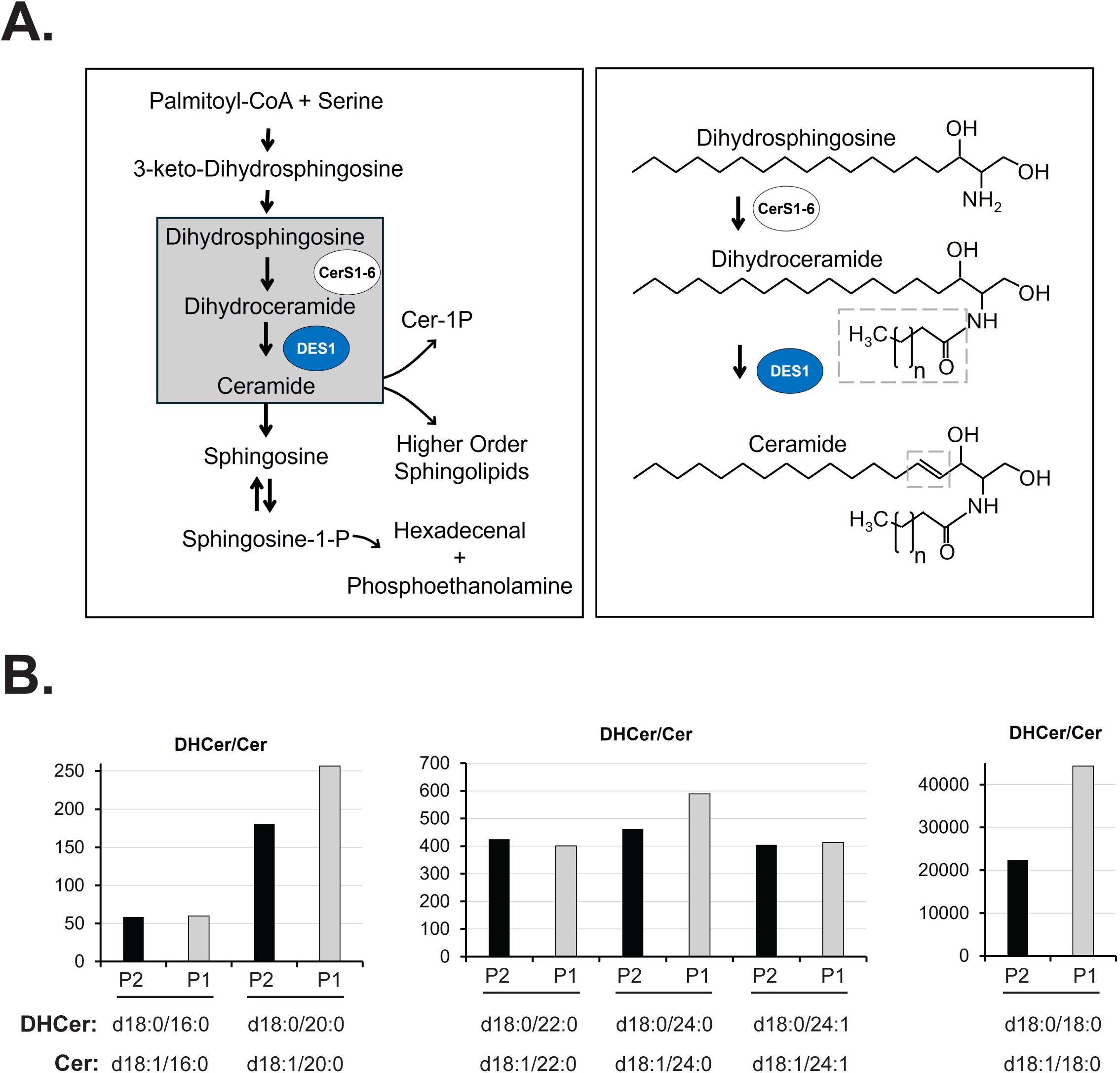
Splice site variant leads to loss-of-function. **(A)** The role of DES1, the product of the *DEGS1* gene, in sphingolipid metabolism. LEFT: Sphingolipid biosynthesis begins with condensation of serine and palmitoylCoA, resulting in 3-keto dihydrosphingosine. This is reduced, forming dihydrosphingosine. Ceramide synthases (CerS) acylate dihydrosphingosine at the free amino group with fatty acids of varying chain lengths and saturation. DES1 desaturates the dihydrosphingosine backbone, introducing a double bond that converts dihydrosphingosine to sphingosine, and correspondingly converts dihydroceramide into ceramide. Formation of all ceramides requires this step. RIGHT: The molecular events catalyzed by CerS and DES1 corresponding to the gray box on the left are shown in detail, with the catalytic activities of both enzymes highlighted in the gray dotted box areas. **(B)** Participants exhibit a profoundly high plasma dihydroceramide to ceramide ratio. Results are shown as fold elevation over mean control ratios, which are arbitrarily set at a value of 1. Participant two, black bars; Participant one, gray bars.

## Discussion

We identified a new homozygous splice site variant in *DEGS1* c.825+4_825+5delAGinsTT, located within the 5’ splice site of *DEGS1* exon two. The variant was present in three participants with HLD18 from two families of the same ancestry who were unrelated at least to the fourth degree. The available parents were heterozygous for the variant and did not exhibit features of HLD18. The variant was absent from population databases, such as gnomAD, dbSNP, TopMed, DiscovEHR, and had not been previously reported in ClinVar. Through RNA sequencing, cell-based splicing assays, RNA structure probing and mass spectrometry analysis, we conclusively showed that the splice site variant was sufficient to induce exon two skipping, and led to the loss of DES1 sphingolipid delta(4)-desaturase protein function. Based on these results, we are submitting the variant to Clinvar as Likely Pathogenic for HLD18.

Interestingly, the findings in participants profiled in this study contrast with a milder disease progression of HLD18 for some individuals reported by Pant et al. (2019), in which acquired microcephaly was present in 3/19, independent sitting attained by 8/19, and contractures present in only 2/19. For full description of clinical findings for the participants in this study and others with HLD18, see Supplemental Table 3.

Quantification of a diverse range of sphingolipids revealed a clear pattern consistent with and pathognomonic of *DEGS1* loss-of-function, accumulation of substrates and paucity of products.^4^ The combination of extremely reduced Cers and elevated DhCers cannot readily be explained by any other enzymopathy or disease state.

Successful splice-modulating ASOs like Spinraza follow a logic to mask splicing silencers to rescue splicing.^35,36^ Our conventional ASO walk did not identify splice-modulating ASOs for the pathogenic variant, largely because the molecular mechanism by which *DEGS1* c.825+4_825+5delAGinsTT affects splicing is complex and involves RNA structure changes.

Our ASO walk results are consistent with findings that large internal exons, like DEGS1 exon two, contain a high density of enhancers that are important for splicing fidelity.^37^ Taken together with our RNA folding models, the ASO walk data suggested that splicing regulatory elements may become inaccessible in the context of the *DEGS1* variant. This complicated splicing abnormality highlights the need for exploring more ASO design strategies, including: oligo length optimization, target specificity, and specific chemical modifications.^38^

DNA sequencing-based genetic testing of children with a suspected leukodystrophy has a relatively high yield of specific diagnoses.^39^ However, this high-throughput testing modality gives rise to numerous variants of uncertain significance.^40,41^ While there are examples of RNA sequencing,^42^ splicing assays,^8^ and metabolic profiling^4,43^ being used to establish the pathogenicity of VUSs in individuals with leukodystrophies, these approaches are currently underutilized.

In this study, we utilized a combination of RNA and protein studies to conclusively demonstrate that a 5’ splice site mutation of *DEGS1* exon two is pathogenic, highlighting the collaborative interdisciplinary approach that was needed to resolve a VUS and establish the molecular mechanisms of disease. Given the high rate of VUSs, as genomic sequencing becomes more common in clinical practice, we must establish cost-effective and scalable frameworks for their resolution using multi-disciplinary approaches.

## Supporting information

VariantValidator_HGVS Nomenclature

## Data Availability

Sequencing data has been uploaded to the Analysis Visualization and Informatics Lab-(AnVIL) at the National Human Genome Research Institute. Clinical data is available from the authors on reasonable request.

## Acknowledgements

We would like to thank the participants and their families for their invaluable participation.

## Author Contributions

**Conceptualization:** A.J.F., A.N.B., A.S., O.M.V.; **Data curation:** A.S., J.D.S., J.Y.L.; **Formal analysis:** A.C., B.S.-J., H.C.B., J.A., J.D.S., J.Y.L., N.R., O.M.V., P.M., U.H., V.T., Y.M.; **Funding acquisition:** A.N.B., A.S., J.D.S., J.R.S., M.D.S., N.R., O.M.V., T.Y.; **Investigation:** A.J.F., A.S., B.S.-J., G.C., H.C.B., J.A., J.D.S., J.R.S., J.S., J.T., J.Y.L., M.D.S., M.G., P.D., P.M., T.Y., U.H., V.T., Y.M.; **Methodology:** D.R.M., G.C., H.C.B., J.A., J.D.S., J.R.S., J.Y.L., M.D.S., V.T.; **Project administration:** H.C.B., V.T.; **Resources:** J.D.S., N.R.; **Software:** D.R.M., H.C.B., J.A.; **Supervision:** A.J.F., A.N.B., H.C.B., J.D.S., J.R.S., J.V.Z, N.R., P.D., V.T.; **Validation:** G.C., H.C.B., M.D.S., M.G., V.T.; **Visualization:** G.C., H.C.B., J.A., M.D.S., V.T.; **Writing-original draft:** H.C.B., J.D.S., J.Y.L., V.T.; **Writing-review & editing:** A.C., A.J.F., A.N.B., A.S., B.S.-J., D.R.M., G.C., H.C.B., J.A., J.D.S., J.R.S., J.S., J.T., J.V.Z, J.Y.L., M.D.S., M.G., N.R., O.M.V., P.D., P.M., T.Y., U.H., V.T., Y.M.

## Ethics Declaration

Written, informed consent was obtained using a consent form from the [REDACTED] study that was approved by the Institutional Review Board (IRB) at [REDACTED].

---

**Supplemental Figure 1.**
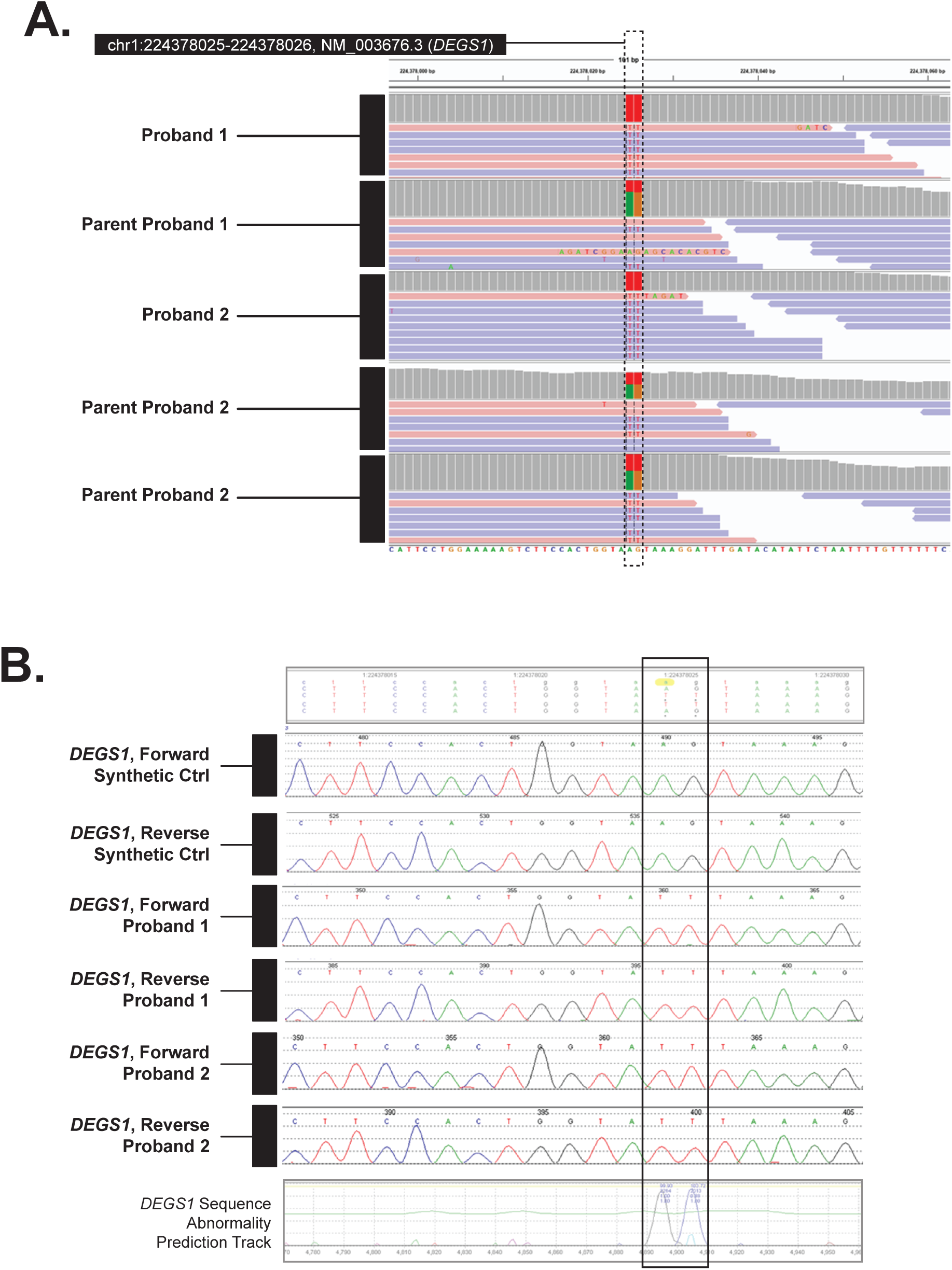
Homozygous non-canonical *DEGS1* splice site variant detected in two participants with hypomyelinating leukodystrophy 18. **(A)** Integrative Gene Viewer plots from exome sequencing showing homozygous variants in two participants and heterozygous variants in available parents. The variants map to the 5’ splice site of exon two from the *DEGS1* gene (NM_003676, chromosomal position chr1:224378025-224378026 on hg19). The position of the variant is boxed. **(B)** Sanger sequencing confirming GRCh38NC_000001.11:g.224190323_224190324delinsTT to be homozygous in participants one and two.

**Supplemental Figure 2.**
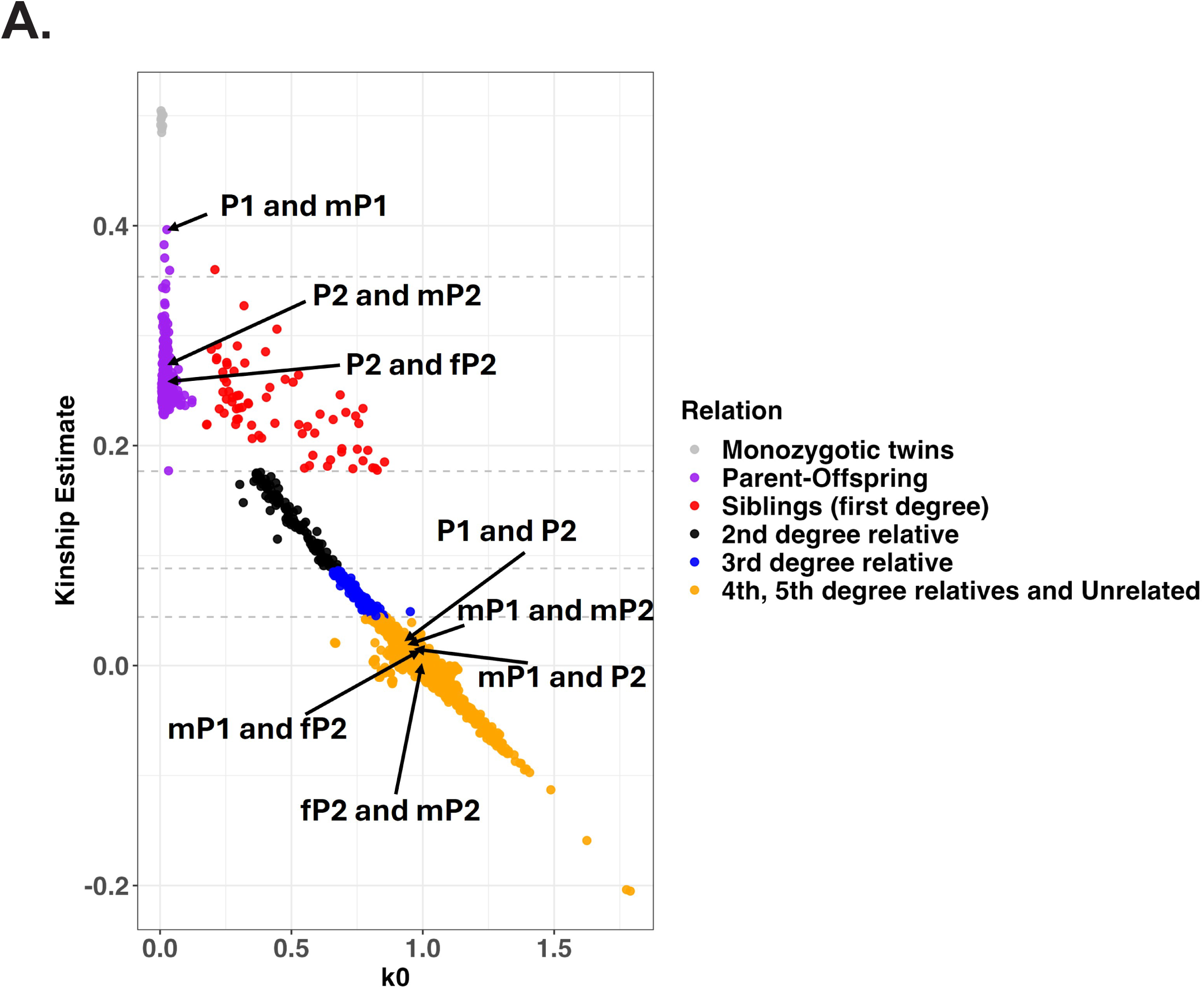
Relationships among participants in the [REDACTED] study. Each dot represents a pair of individuals among the sequenced [REDACTED] participants and parents. The Y axis shows the kinship coefficient, which measures how genetically similar two individuals are. Higher values mean the individuals are more closely related. The x-axis (k0) shows the chance that any shared allele between two individuals is identical by state (IBS). IBS means the alleles are the same, whether they came from a common ancestor or not. P1 and P2 represent participants one and two respectively. Female and male parents of the participants are represented by fP and mP, respectively.

**Supplemental Figure 3.**
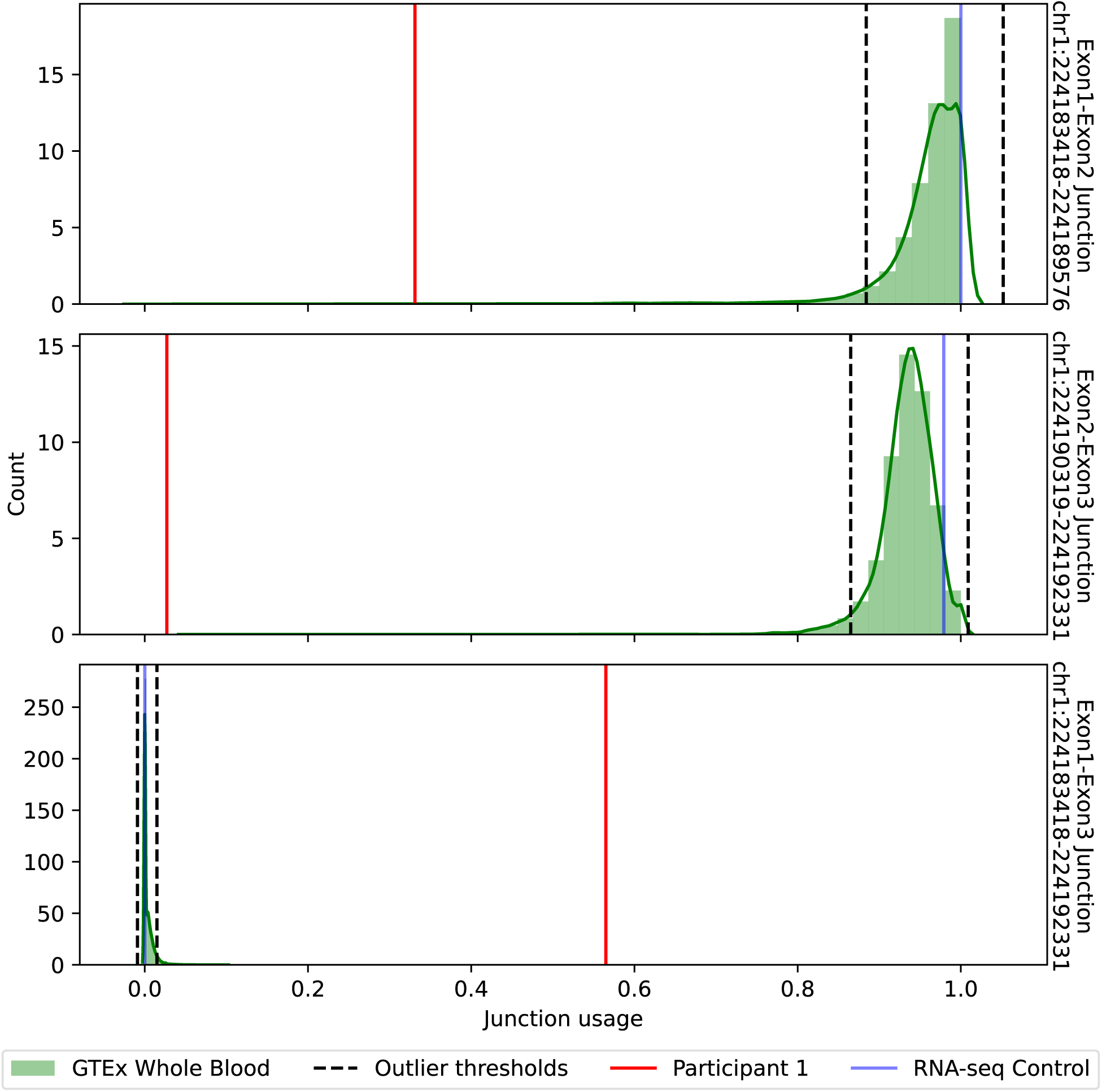
Junction usage in participant one is exceptional relative to the control sample and whole blood in GTEX. Participant one has exceptionally low usage of commonly used *DEGS1* junctions (top two panels), far below the outlier thresholds defined by junction usage in whole blood in GTEx. Usage of the *DEGS1* exon 1-3 junction is extremely rare in the control sample and GTEx whole blood, but abundant in Participant one. Genome coordinates are for hg38.

**Supplemental Figure 4.**
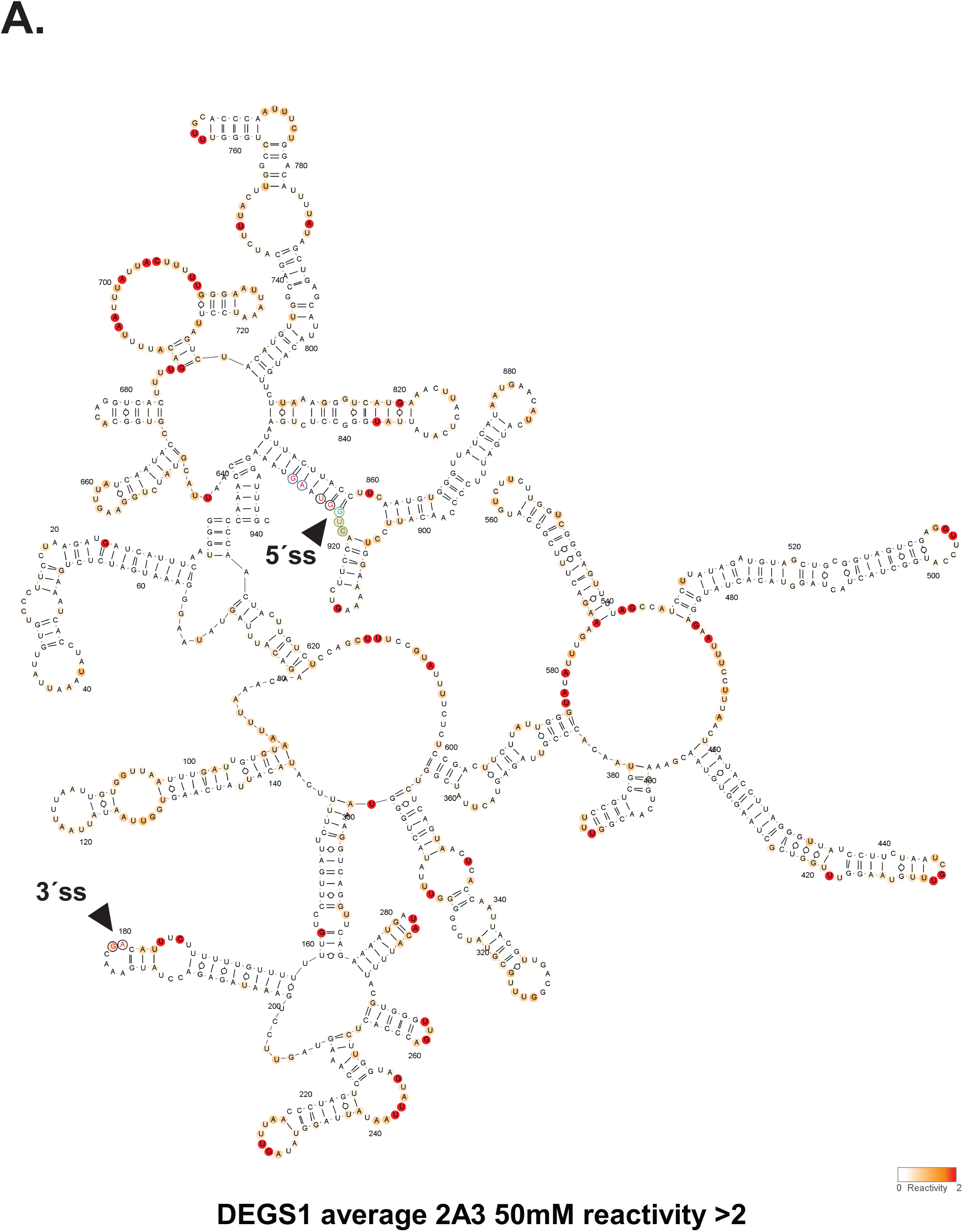
Two-dimensional RNA structure prediction for the reference context of *DEGS1* exon two. SHAPE-derived secondary structural model for *DEGS1* exon two reference and flanking intron sequences, as assayed in splicing reporter assays. Bases are colored according to their normalized 2A3 SHAPE reactivity. All nucleotide position numbering shown is based on the IVT RNA template used for SHAPE probing, from the 5′ to 3′ orientation. Splice sites are indicated with black arrows and appropriate text.

**Supplemental Figure 5.**
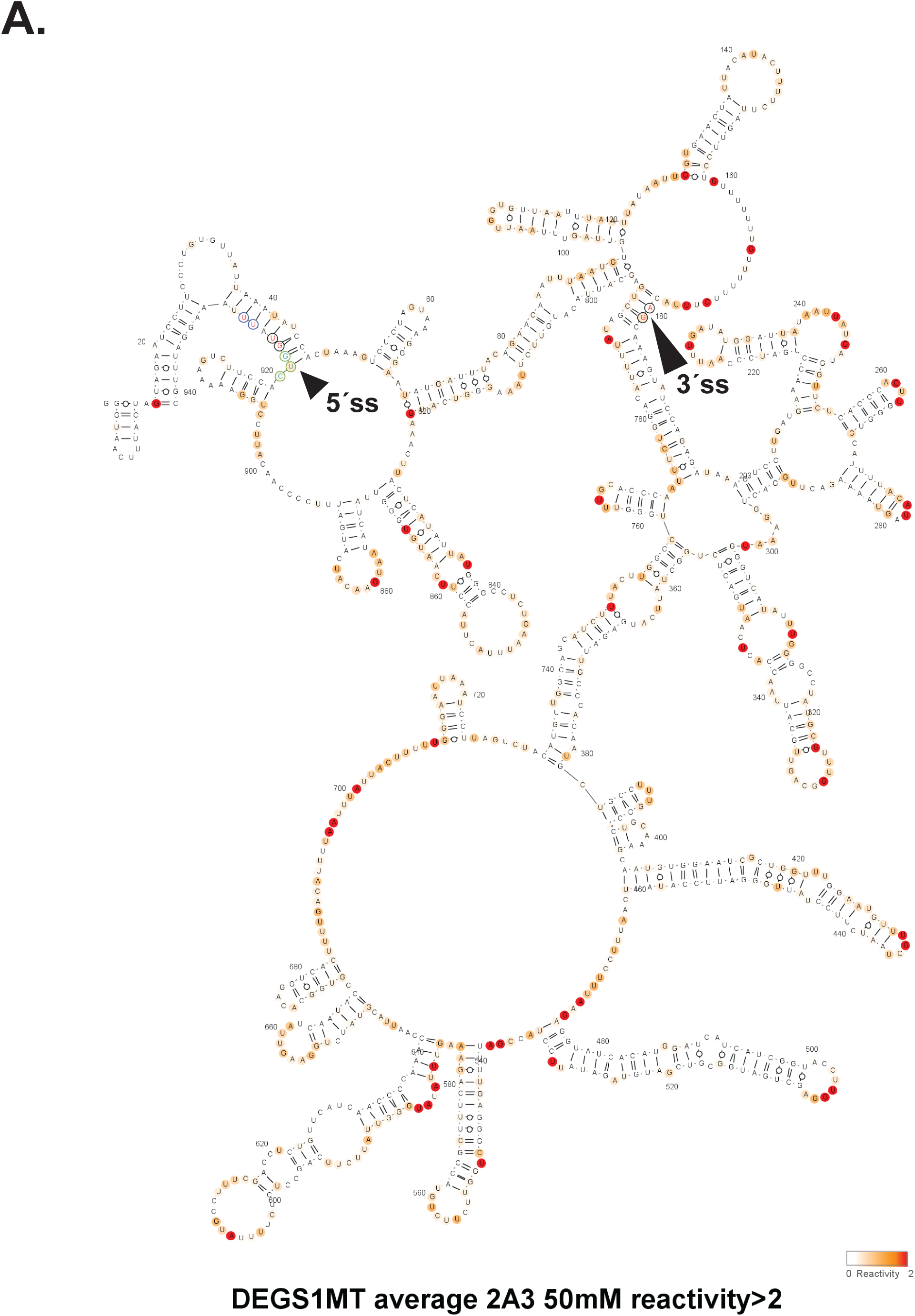
Two-dimensional RNA structure prediction for the splice site variant context of *DEGS1* exon two. SHAPE-derived secondary structural model for *DEGS1* exon two variant and flanking intron sequences, as assayed in splicing reporter assays. Bases are colored according to their normalized 2A3 SHAPE reactivity. All nucleotide position numbering shown is based on the IVT RNA template used for SHAPE probing, from the 5′ to 3′ orientation. Splice sites are indicated with black arrows and appropriate text.

**Supplemental Figure 6.**
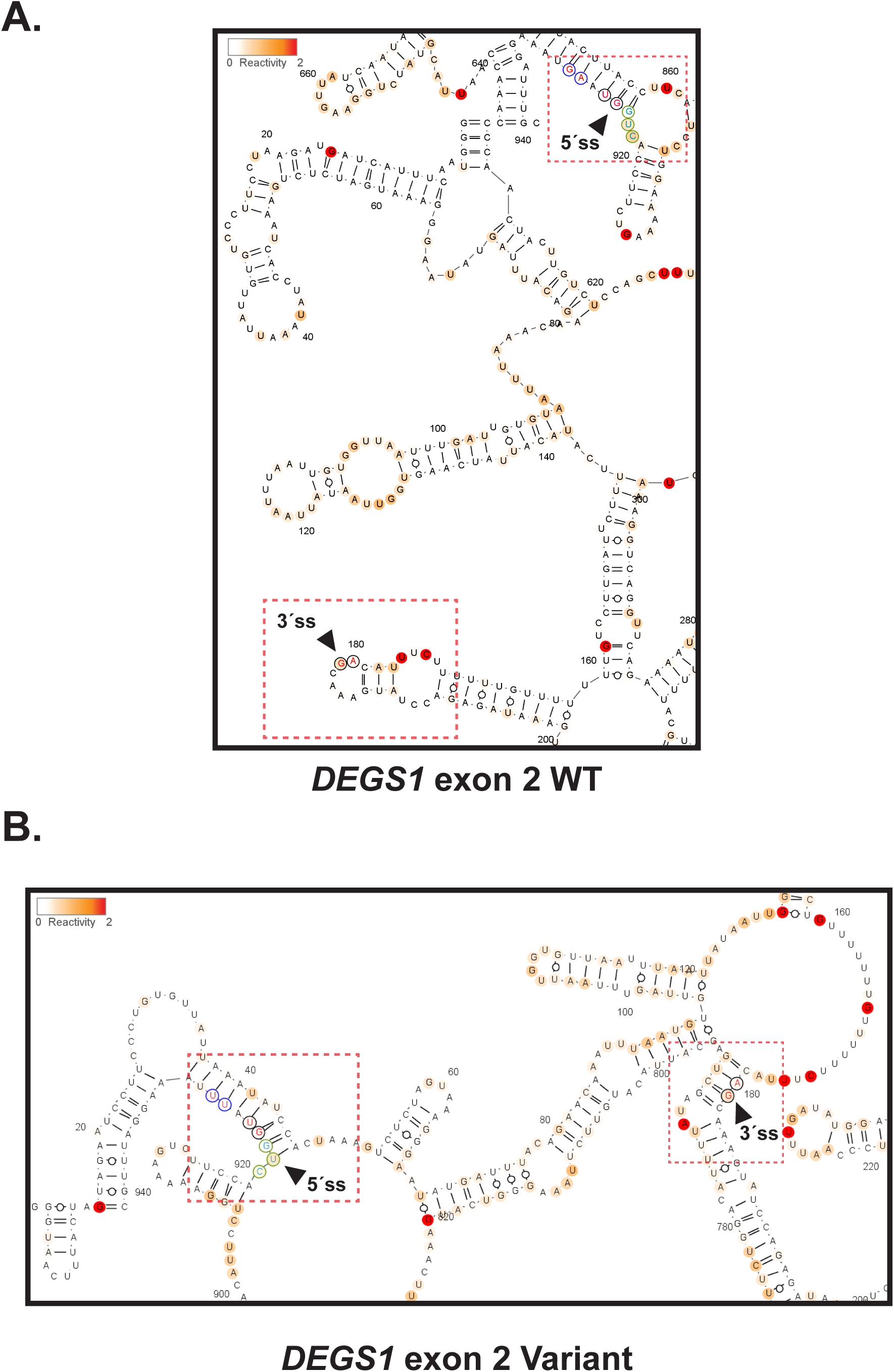
Examining the structural accessibility of splice sites between the reference and variant context of *DEGS1* exon two. The figure shows the splice sites for the reference (i.e., wildtype; WT), as shown in Panel **(A)**, and for the variant, as shown in Panel **(B)**. Respective splice sites are indicated with a black arrow and are also boxed in red.

**Supplemental Figure 7.**
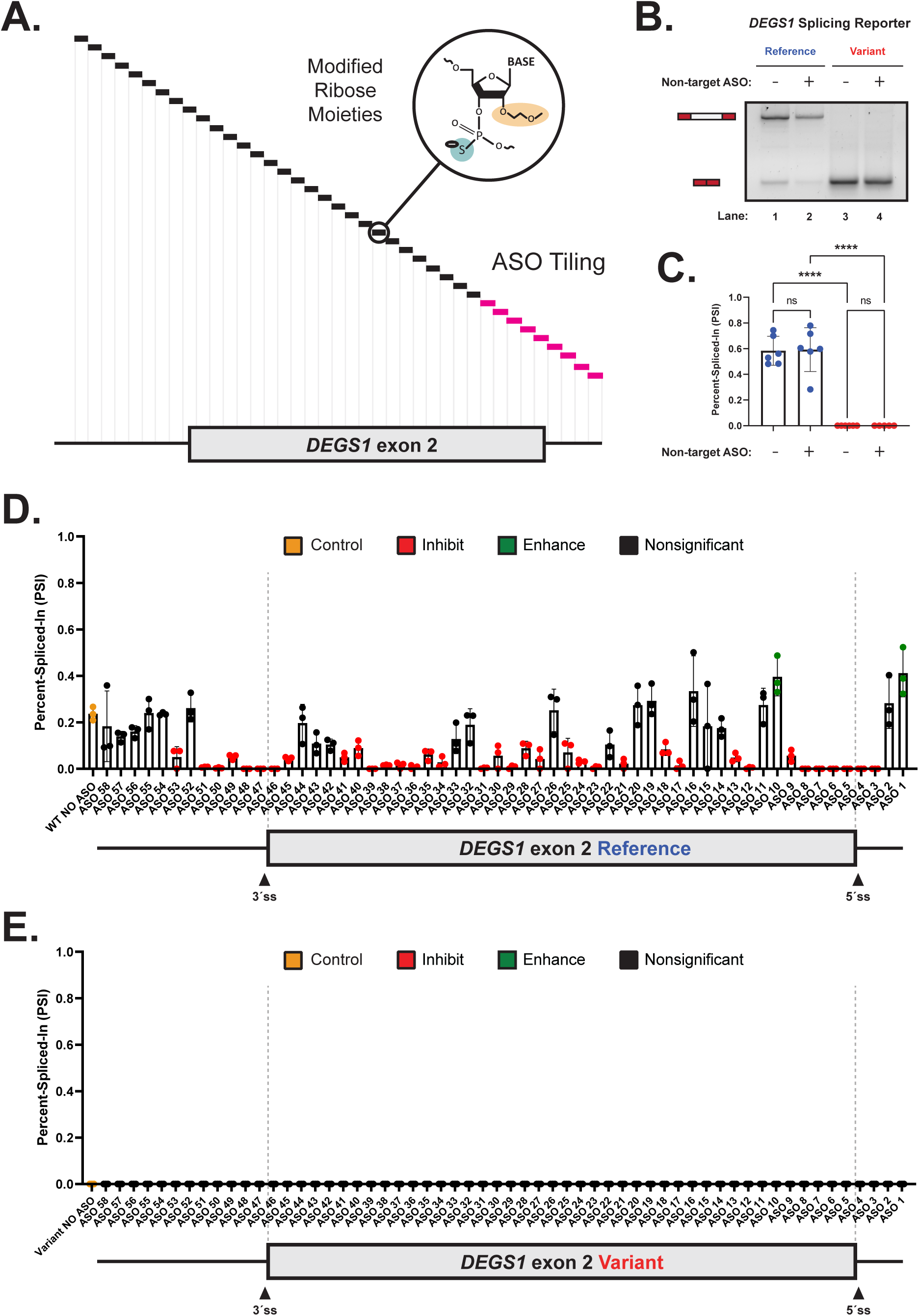
*DEGS1* exon two is highly dependent on exonic splicing enhancers for its splicing. **(A)** A mock schematic of the antisense oligonucleotide (ASO) walk conducted in this study. All ASOs tile across *DEGS1* exon two and its flanking introns. Each ASO used in our walks was 18 nucleotides in length and was designed using ribose sugars that were modified with a 2′-methoxyethyl group (2′-MOE, highlighted in light orange), and the phosphate backbone was modified to a phosphorothioate backbone (highlighted in light blue). Black boxes represent 18-mer ASOs that were contiguous by design, whereas hot-pink boxes represent 18-mer ASOs that had 10 nt overlaps between the preceding and proceeding ASO. **(B)** A representative agarose gel demonstrated no significant difference between conditions with and without a non-targeting ASO being co-transfected with our wildtype (WT; blue) or variant (red) splicing reporter. Expected mRNA isoforms including or excluding *DEGS1* exon two are also annotated to the left of the agarose gel. **(C)** Percent-Spliced-In (PSI) plot quantifying the non-targeting ASO’s impact on *DEGS1* exon two splicing as shown in (B). **(D, E)** PSI plots quantifying our ASO walk data on the sequence context corresponding to the WT or variant of *DEGS1* exon two, respectively. A schematic model of *DEGS1* and its flanking introns is shown at the bottom of each PSI plot to illustrate relative positions of ASOs, and for which sequence context. ASOs that significantly inhibited splicing are indicated in red, whereas those that significantly enhanced splicing are indicated in green. The control ASO is depicted in yellow, and non-significant ASOs are depicted in black. Statistical significance between comparisons shown is denoted by asterisks (i.e., ****) that represent a *P* ≤ 0.0001. Statistical significance was determined using analysis of variance (ANOVA), and Dunett’s post-hoc test. Each condition tested and presented in this figure contains a minimum of three independent/biological replicates.

**Supplemental Figure 8.**
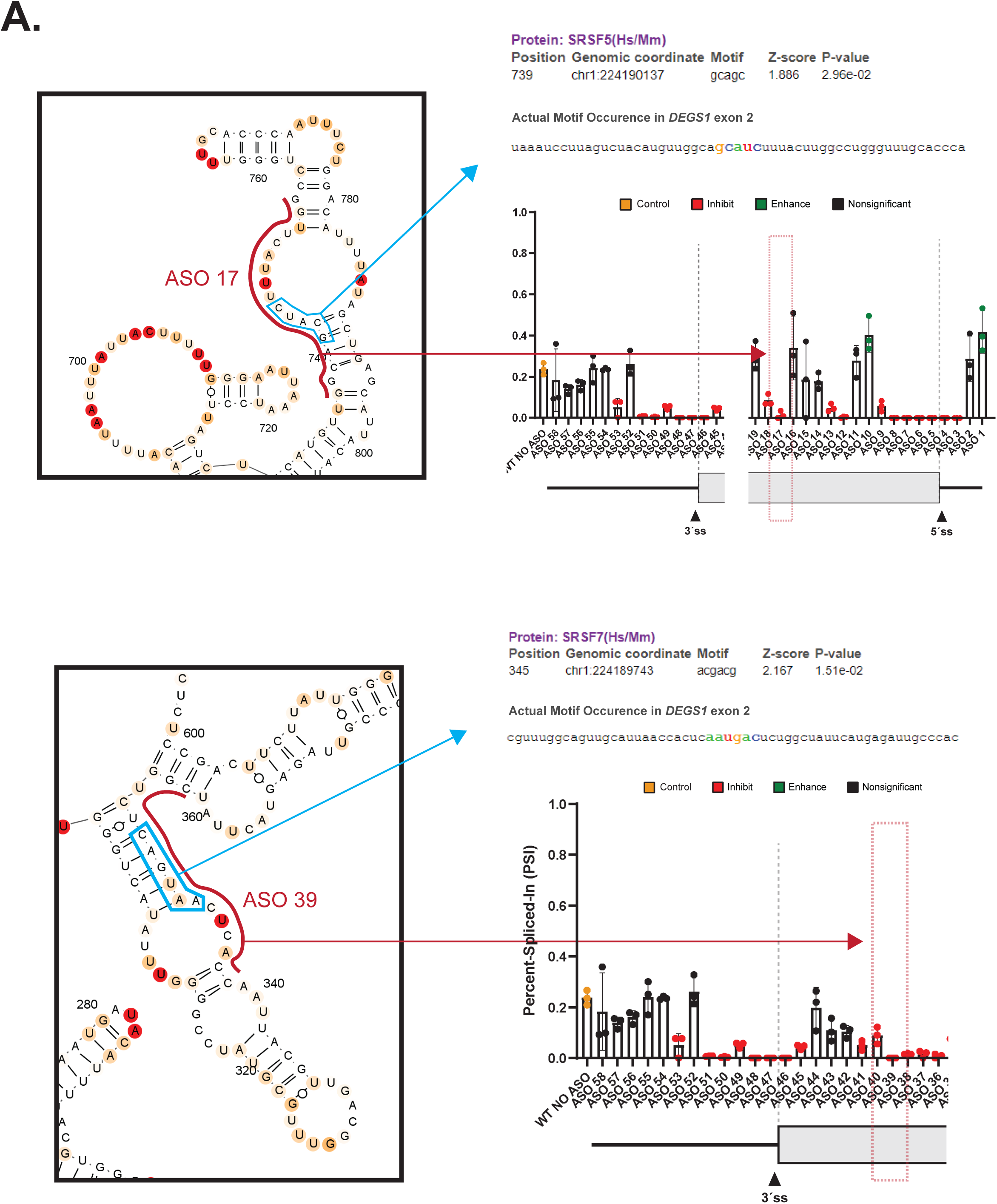
Cross-referencing RBPmap predictions to *DEGS1* exon two SHAPE and ASO data. Among a list of predictions, for example, RBPmap predicts two putative binding sites for splicing factors known to enhance splicing. The splicing factor implicated is described, showing their motif confidence score and their position within our sequence context assayed, as indicated following the light blue arrow. The effect an ASO has in interfering with these putative splicing enhancers are indicated with red annotations such as text, arrows, and dashed boxes.

**Supplemental Figure 9.**
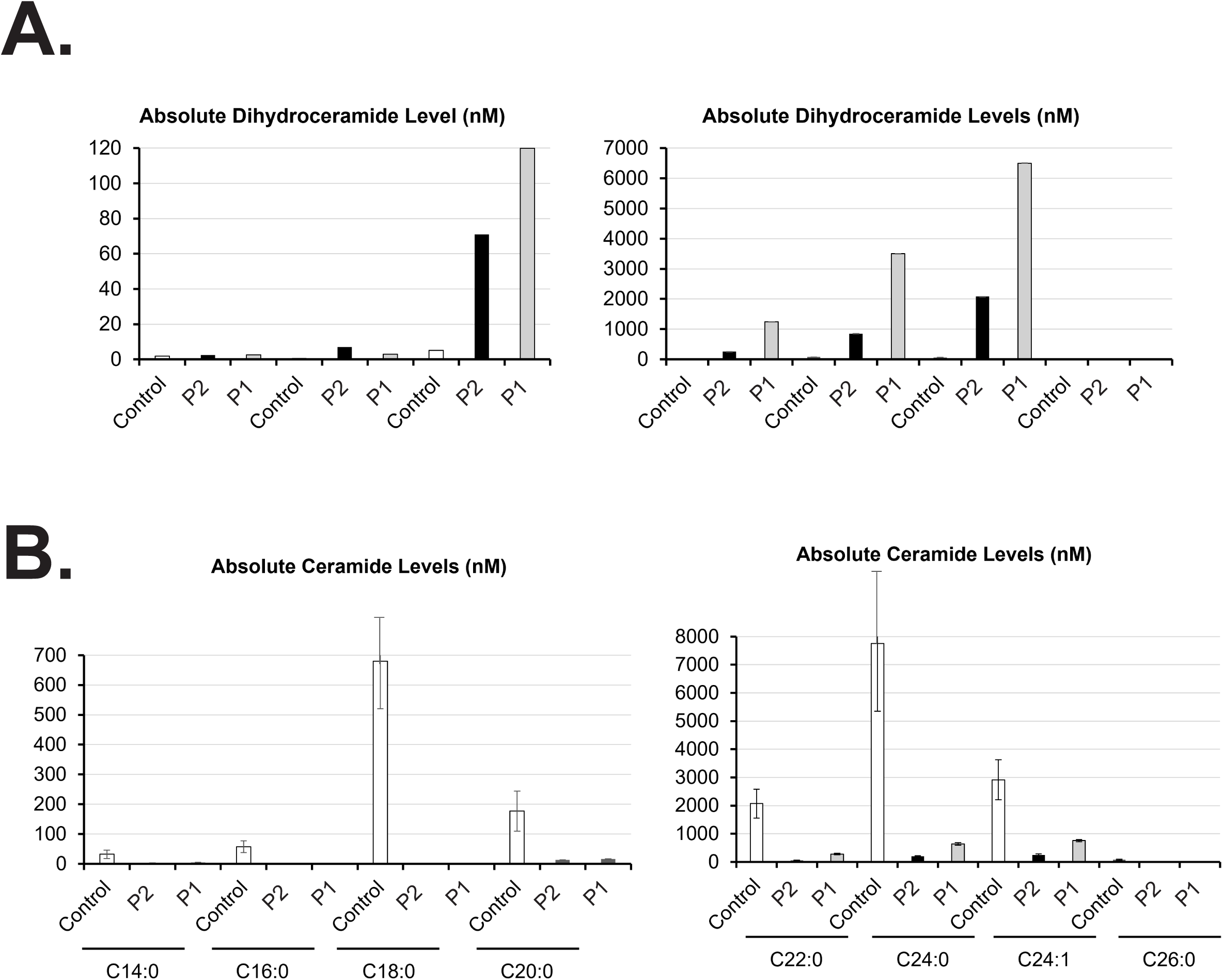
Participants exhibit high levels of dihydroceramides and low levels of ceramides in plasma. **(A)** Two patients with homozygous *DEGS1* splice site variants (Participant one (P1) and participant two (P2)) exhibit profound accumulation of the *DEGS1* dihydroceramide substrates in their plasma compared to healthy pediatric controls (n=9). Shown are results for seven dihydroceramide species with fatty acids of the following chain length and saturation (C16:0, C18:0, C20:0, C22:0, C24:0, C24:1, C26:0). **(B)** Two patients with homozygous *DEGS1* variants (P1, P2) exhibit profound deficiency of the *DEGS1* ceramide products in their plasma compared to healthy pediatric controls (n=9). Shown are results for eight ceramide species with fatty acids of the following chain length and saturation (C14:0, C16:0, C18:0, C20:0, C22:0, C24:0, C24:1, C26:0). Results from P2 and P1 are represented by black bars and gray bars, respectively. Results from a control participant are represented by white bars.

