## Supplementary material for "A novel splice site variant in *DEGS1* leads to aberrant splicing and loss of DEGS1 enzyme activity, a VUS resolved": VariantValidator_HGVS Nomenclature

### Submitted Variant

NM_003676.4:c.825+4_825+5delinsTT

- Selected genome build: GRCh37
- Map location: 1q42.11
- Transcript Flag: MANE Select
- CCDS ID: CCDS1540.1

### Versions

- VariantValidator 2.2.1.dev691+g8946f76
- vv_hgvs 2.2.0
- VVDb vvdb_2024_5
- VVTA vvta_2024_01
- VVSeqRepo VV_SR_2024_01/master

### Recommended Variant Descriptions

1. HGVS guidelines recommend using genomic and transcript descriptions in all publications
2. Use of the three- or one-letter amino acid alphabet is optional, but three-letter is recommended

### Genomic descriptions

| Reference Sequence Type | Variant Description |
|---|---|
| Chromosomal GRCh37 | NC_000001.10:g.224378025_224378026delinsTT |
| Chromosomal GRCh38 | NC_000001.11:g.224190323_224190324delinsTT |

### Transcript and protein descriptions

| Reference Sequence Type | Variant Description |
|---|---|
| Transcript | NC_000001.10(NM_003676.4):c.825+4_825+5delinsTT |
| Protein three letter code | NP_003667.1:p.? |
| Protein single letter code | NP_003667.1:p.? |

### Gene Information

| Attribute | Identifier | Source |
|---|---|---|
| Symbol | DEGS1 | HGNC |
| Name | delta 4-desaturase, sphingolipid 1 | HGNC |
| HGNC ID | HGNC:13709 | HGNC |

MANCHESTER 1824
The University of Manchester

Copyright © 2016-2024 VariantValidator Contributors

UNIVERSITY OF LEICESTER
